# Causal relationships between risk of venous thromboembolism and 18 cancers: a bidirectional Mendelian randomisation analysis

**DOI:** 10.1101/2023.05.16.23289792

**Authors:** Naomi Cornish, Philip Haycock, Hermann Brenner, Jane C. Figueiredo, Tessel Galesloot, Robert C Grant, InterLymph consortium, INVENT-MVP consortium, Mattias Johansson, Daniela Mariosa, James McKay, Rish Pai, Andrew J Pellatt, Jewel Samadder N., Jianxin Shi, Florian Thibord, David-Alexandre Trégouët, Catherine Voegele, Chrissie Thirlwell, Andrew Mumford, Ryan Langdon

## Abstract

**Background:** People with cancer experience high rates of venous thromboembolism (VTE). Additionally, risk of subsequent cancer is increased in people experiencing their first VTE. The causal mechanisms underlying this association are not completely understood, and it is unknown whether VTE is itself a risk factor for cancer.

**Methods:** We used data from large genome-wide association study meta-analyses to perform bi-directional Mendelian randomisation analyses to estimate causal associations between genetically-proxied lifetime risk of VTE and risk of 18 different cancers.

**Results:** We found no conclusive evidence that genetically-proxied lifetime risk of VTE was causally associated with an increased incidence of cancer, or vice-versa. We observed an association between VTE and pancreatic cancer risk (odds ratio for pancreatic cancer 1.23 (95% confidence interval 1.08 - 1.40) per log-odds increase in risk of VTE, *P =* 0.002). However, sensitivity analyses revealed this association was predominantly driven by a variant proxying non-O blood group, with inadequate evidence from Mendelian randomisation to suggest a causal relationship.

**Conclusions:** These findings do not support the hypothesis that genetically-proxied lifetime risk of VTE is a cause of cancer. Existing observational epidemiological associations between VTE and cancer are therefore more likely to be driven by pathophysiological changes which occur in the setting of active cancer and anti-cancer treatments. Further work is required to explore and synthesise evidence for these mechanisms.

**Key messages:**

1) There is strong observational evidence that active cancer is associated with venous thromboembolism.
2) It is currently unknown whether venous thromboembolism is a risk factor for cancer.
3) We applied a bi-directional Mendelian randomisation framework to appraise the causal relationships between genetically-proxied risk of venous thromboembolism and 18 different cancers.
4) Overall, there was no clear evidence from Mendelian randomisation that lifetime-elevated risk of venous thromboembolism is causally associated with an increased risk of cancer, or visa versa.

## Introduction

Venous thromboembolism (VTE), which includes deep vein thrombosis and pulmonary embolism, is the third most common cause of death from cardiovascular disease globally.[1] Over 20% of all VTE events occur in people with pre-existing cancer, for whom the relative risk of VTE is at least 5 times higher than age-matched non-cancer controls.[2] Evidence from in-vitro and animal models shows that active cancer can precipitate a prothrombotic state; many tumours directly activate platelets, produce procoagulant proteins such as tissue-factor, or alter the vascular endothelium, all of which may increase the risk of thrombosis.[3] Systemic anti-cancer therapy or surgery and their resultant complications, including sepsis and hospitalisation, are also powerful risk-factors for VTE.[2]

It is currently unknown whether a predisposition to VTE is causally associated with cancer incidence. Over 5% of people presenting with a first VTE are subsequently diagnosed with cancer within the ensuing year[4,5] and several studies have indicated that cancer risk may be elevated over the longer term for people with a history of VTE.[6–9] Experiments in mice indicate that pro-thrombotic proteins, including tissue factor and fibrinogen, facilitate tumour growth, survival and metastasis.[10,11] However, observational studies examining whether treatment with antiplatelet or anticoagulant medication reduces risks of cancer have shown conflicting results.[12–14]

Attempts to elucidate complex causal relationships between VTE and cancer using traditional observational studies are complicated by difficulties in ascertaining direction of causality, in addition to being susceptible to unmeasured and residual confounding from risk factors which are common to both VTE and cancer, including smoking, obesity and co-existing inflammatory conditions.[2] Mendelian randomisation (MR) analysis addresses some of these limitations. It employs genetic variants, typically single nucleotide polymorphisms (SNPs), as instrumental variables (IVs) to proxy the effect of an exposure on an outcome. As SNPs are randomly allocated and fixed at conception, they are unconfounded by acquired and environmental risk factors.[15] Here we apply a bi-directional MR approach to examine the effect of genetically-proxied VTE risk on the risk of 18 cancers, and conversely the effect of genetically-proxied cancer-risk on the risk of VTE.

## Methods

### ii) Data sources

We obtained European-ancestry summary genetic data from large consortia-driven genome-wide association study (GWAS) meta-analyses examining risk of VTE and 18 common cancers, respectively (**Table 1**). Summary genetic data included the effect size of each SNP (log-odds ratio), standard-error of the effect size, *P*-value, sample size (including case-control ratio) and, where available, the effect allele frequency in the GWAS study population.

**Table 1.** Source of GWAS data used for bi-directional Mendelian randomisation analysis.

| Trait | Study, date | n cases | n controls | Sample size |
| --- | --- | --- | --- | --- |
| Venous thromboembolism | Thibord *et al*, 2022[16] | 71 771 | 1 059 740 | 1 131 511 |
| Breast cancer | Zhang *et al*, 2020[17] | 133 384 | 113 789 | 247 173 |
| Prostate cancer | Schumacher *et al*, 2018[18] | 79 194 | 61 112 | 140 306 |
| Endometrial cancer | O'Mara *et al*, 2018[19] | 12 906 | 108 979 | 121 885 |
| Colorectal cancer | Huyghe *et al*, 2019[20] | 55 168 | 65 160 | 120 328 |
| Melanoma | Landi *et al*, 2020[21] | 30 134 | 81 415 | 111 549 |
| Lung Cancer | McKay *et al*, 2017[22] | 29 266 | 56 450 | 85 716 |
| Ovarian cancer | Phelan *et al*, 2017[23] | 25 509 | 40 941 | 66 450 |
| Kidney Cancer | Scelo *et al*, 2017[24] | 10 784 | 20 406 | 31 190 |
| Oesophageal cancer | Gharahkhani *et al*, 2016[25] | 4112 | 17 159 | 21 271 |
| Pancreatic cancer | PanScan/PanC4, 2022<sup>a</sup> | 9055 | 7203 | 16 258 |
| Diffuse large B cell lymphoma | Cerhan *et al*, 2014[26] | 3857 | 7666 | 11 523 |
| Chronic lymphocytic leukaemia | Berndt *et al*, 2013[27] | 3100 | 7667 | 10 767 |
| Follicular lymphoma | Skibola *et al*, 2014[28] | 2728 | 7758 | 10 468 |
| Oral cancer | Lesseur *et al*, 2016[29] | 2700 | 5984 | 8684 |
| Oropharyngeal cancer | Lesseur *et al*, 2016[29] | 2433 | 5984 | 8417 |
| Glioma | Melin *et al*, 2017[30] | 4572 | 3286 | 7858 |
| Marginal zone lymphoma | Vijai *et al*, 2015[31] | 825 | 6221 | 7046 |
| Bladder cancer | Nijmegen Bladder Cancer Study<sup>a</sup> | 1799 | 4745 | 6544 |

<sup>a</sup>Indicates unpublished data

To examine the association between genetically-proxied risk of VTE and each cancer, we extracted risk SNPs associated with VTE at *P*<5×10^-8^ from a VTE GWAS conducted by Thibord *et al*.[32] We clumped SNPs using a strict linkage disequilibrium (LD) threshold of r^2^<0.001 (10,000kb sliding window) to ensure independence. Clumping was performed with the ‘TwoSampleMR’ R package[33] using European-ancestry reference panels from the 1000 Genomes Project.[34] If exposure SNPs were absent from the reference panel, these were excluded from the analysis.

Summary statistics for the SNPs in our VTE IV were then extracted from each cancer risk GWAS. If a VTE-risk SNP was not present in the GWAS summary statistics for a given cancer, an alternative SNP in high LD with the target SNP (r^2^ ý0.8) was used as a proxy (if available). We harmonised exposure and outcome data to ensure that effect estimates corresponded to the same allele for each SNP across the VTE and cancer datasets. Coding-strand ambiguities for palindromic SNPs were resolved using effect allele frequencies if possible; palindromic SNPs with intermediate effect allele frequencies (0.42 – 0.58) were excluded.[33] Effect allele frequencies were not available for the oesophageal cancer and glioma GWAS. For oesophageal cancer we confirmed the coding-strand with study authors to facilitate data-harmonisation; for glioma all palindromic SNPs were excluded.

To perform the analysis in the opposite direction (with genetic risk of cancer as an exposure and VTE as an outcome), we used the same process and thresholds described above to select independent GWAS-significant risk SNPs for each cancer from the relevant cancer GWAS, then looked up summary statistics for the cancer-risk SNPs in the VTE GWAS.

### ii) Statistical analyses

Causal estimates from MR are underpinned by three core assumptions: 1) the genetic variants which are used as IVs are strongly associated with the exposure; 2) there are no confounders of the genotype-outcome relationship; 3) the genetic variants affect the outcome only via the exposure and not through an alternative pathway (violation of this is known as ‘horizontal pleiotropy’).[35]

We assessed the strength of each SNP-exposure association using F-statistics. We performed Steiger filtering to exclude IVs which explained more variance in the outcome than the exposure.[36] This reduces the risk of using invalid instruments which could impact the outcome via horizontal pleiotropy, or which proxy a reverse-causal pathway from outcome to exposure.

As recommended by published guidelines,[35] we used an inverse variance-weighted multiplicative random effects MR model (MR-IVW) for the primary analysis, with correction for under-dispersion when only a few SNPs were available for analysis. The MR-IVW result is derived from a linear regression of the SNP-outcome and SNP-exposure associations, with each SNP weighted according to the inverse of the variance of the SNP-outcome effect. We assessed heterogeneity between the individual SNP estimates in the MR-IVW model using Cochran’s Q-statistic. The exception to this was for Marginal zone lymphoma, where only a single variant was available as a proxy, therefore the Wald ratio estimator[15] was used to estimate the causal effect.

Since MR-IVW assumes there is no directional pleiotropy in the MR instruments, we performed a range of sensitivity analyses, including MR-Egger, weighted-median, weighted-mode and leave-one-out analyses to test this assumption.[37]

The VTE GWAS data came from a discovery cohort where some novel variants have not been replicated. Therefore, we also performed a sensitivity analysis where we limited the genetic instruments for VTE to previously replicated loci only,[16] to evaluate for bias resulting from either weak instruments or ‘the winner’s curse’.[38]

Previous studies have reported that two powerful VTE risk variants: Factor V Leiden (rs6025) and Prothrombin G20210A (rs1799963), may be associated with cancer incidence.[39–41] These variants have a prevalence of ∼5% and ∼1% respectively in European populations. Notably, carriers of either of these variants have a VTE risk which is 3-5× higher than those with wild-type alleles.[42] We performed a secondary analysis using MR Wald ratios[15] to examine the association between VTE risk as proxied by these individual SNPs and risk of each cancer.

Results are presented in accordance with STROBE-MR guidelines[43] as the odds ratio (OR) and 95% confidence interval (CI) for each outcome per log-odds increase in the risk of exposure. P values (*P*) have been adjusted for multiple-testing using a false discovery rate correction (*FDR-P*). All analyses were performed in R version 4.0.3 using the ‘TwoSampleMR’ package.[33]

## Results

### ii) Mendelian randomisation analyses of the association between genetically-proxied risk of venous thromboembolism and cancer risk

After selecting independent GWAS-significant VTE-risk SNPs (*P*<5×10^-8^, r^2^:s0.001), there were 73 SNPs available as genetic instruments for VTE. These variants explained approximately 3% of the variance in VTE risk in the VTE GWAS cohort.[16,33,44] The number of instrumental variables varied for each VTE-cancer analysis (**Table 2**), as some VTE SNPs were either unavailable for assessment in the cancer GWAS studies, were excluded by Steiger filtering, or could not be harmonised between the datasets due to coding-strand ambiguities. Summary data for the SNPs used in each analysis is shown in **Supplementary Table S1**.

**Table 2.** Number of genetic instruments for venous thromboembolism (VTE) used for each Mendelian randomisation analysis, associated r^2^ (variance in VTE risk explained) and mean F statistic.

| Outcome GWAS | VTE-SNPs available<sup>a</sup> | SNPs excluded<sup>a</sup> | VTE-SNPs used<sup>a</sup> | $r^2$ for VTE | Mean F statistic |
| --- | --- | --- | --- | --- | --- |
| Breast cancer | 68 | 4 | 64 | 0.031 | 208 |
| Prostate cancer | 66 | 2 | 64 | 0.031 | 210 |
| Endometrial cancer | 73 | 7 | 66 | 0.032 | 205 |
| Colorectal cancer | 68 | 2 | 66 | 0.031 | 204 |
| Melanoma | 68 | 4 | 64 | 0.031 | 209 |
| Lung Cancer | 64 | 4 | 60 | 0.031 | 219 |
| Ovarian cancer | 68 | 6 | 62 | 0.031 | 213 |
| Kidney Cancer | 70 | 9 | 61 | 0.031 | 218 |
| Oesophageal cancer | 70 | 6 | 64 | 0.032 | 210 |
| Pancreatic cancer | 45 | 5 | 40 | 0.023 | 254 |
| Diffuse large B cell lymphoma | 71 | 16 | 55 | 0.030 | 237 |
| Chronic lymphocytic leukaemia | 71 | 23 | 48 | 0.030 | 266 |
| Follicular lymphoma | 71 | 23 | 48 | 0.029 | 264 |
| Oral cancer | 68 | 15 | 53 | 0.030 | 242 |
| Oropharyngeal cancer | 68 | 18 | 50 | 0.029 | 255 |
| Glioma | 69 | 26 | 43 | 0.028 | 283 |
| Marginal zone lymphoma | 71 | 29 | 42 | 0.028 | 292 |
| Bladder cancer | 66 | 26 | 40 | 0.028 | 307 |

<sup>a</sup> For each cancer outcome GWAS, 'VTE-SNPs available' refers to the number of VTE risk-SNPs for which a direct correlate or proxy could be identified in the cancer GWAS study; 'SNPs excluded' refers to the number of VTE-SNPs which could not be harmonised due to coding-strand ambiguities or which were excluded after Steiger filtering. 'VTE-SNPs used' refers to the final number of genetic instruments for VTE used in each analysis.

We estimated the OR for each cancer per log-odds increase in genetically-proxied risk of VTE using MR-IVW analysis (**Figure 1**). Increased risk of VTE was associated with an increased risk of pancreatic cancer (OR 1.23 [95% CI, 1.08-1.40], *P*=0.002, *FDR-P=*0.05). A much weaker association in the same direction was seen for ovarian cancer (OR 1.05 [95% CI, 1.00-1.11], *P*=0.04, *FDR-P=*0.29) and endometrial cancer (OR 1.06 [95% CI, 1.00-1.12], *P*=0.05, *FDR-P=*0.31). Sensitivity analyses showed inconsistent estimates of effect between the MR-IVW, MR-Egger, weighted-median and weighted-mode estimates for pancreatic cancer (**Figure 2A, Supplementary Table S2**). There was significant heterogeneity in the VTE IV estimates for pancreatic, ovarian and endometrial cancer as assessed by Cochrane’s Q-statistic. Graphical assessment of the leave-one out plots, single SNP plots and funnel plots (**Supplementary Figures S1-S3**) identified an outlying SNP (rs687289). Removal of this SNP from the analysis virtually abolished the association between VTE and pancreatic, ovarian and endometrial cancer (**Figure 2**).

**Figure 1.**
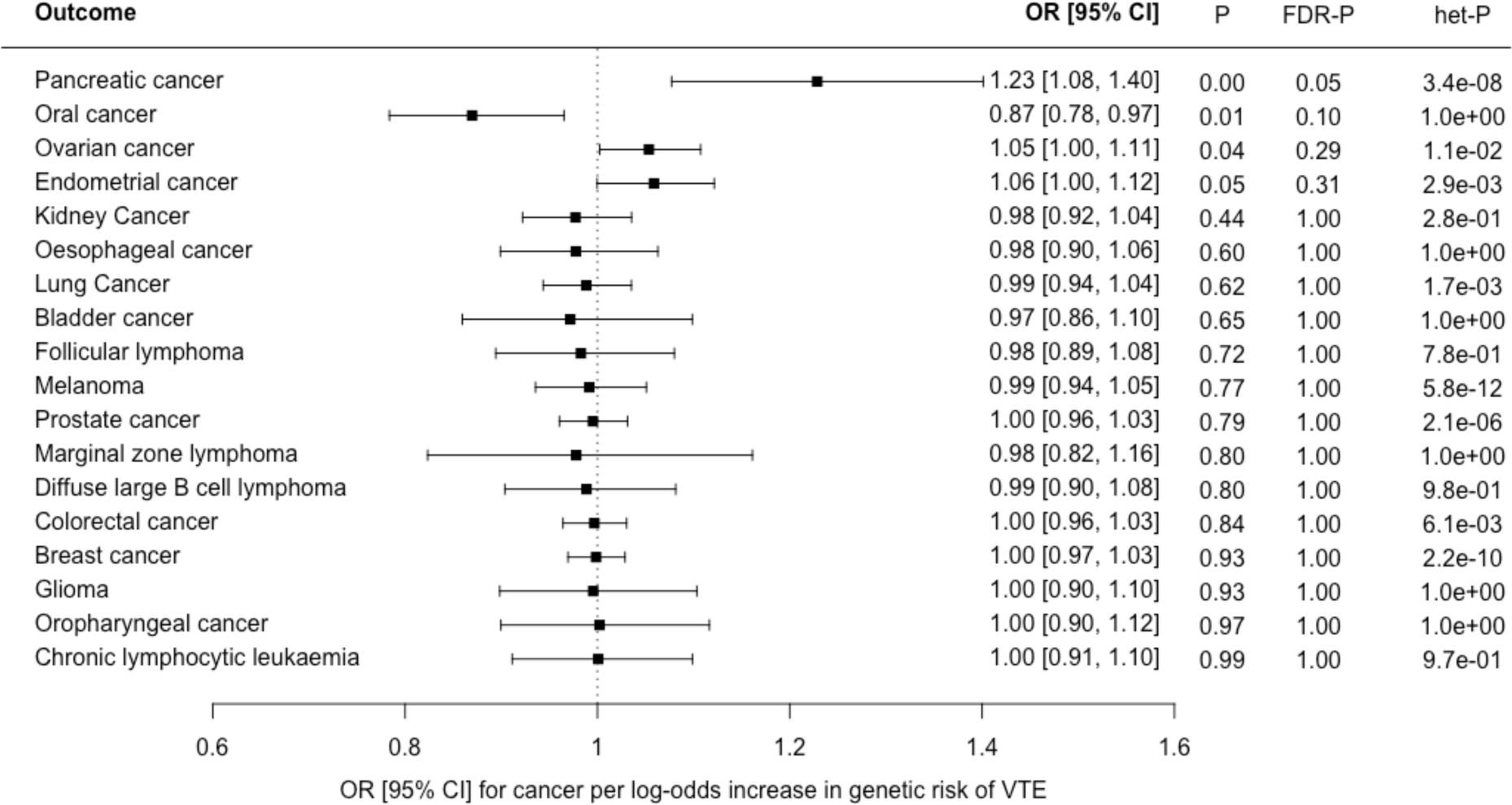
Forest plot showing estimates from Mendelian randomisation inverse variance weighted estimates (MR-IVW) of the effect of genetically-proxied risk of venous thromboembolism (VTE) as an exposure on 18 cancers as outcomes: results are represented as the odds ratio (OR) and 95% confidence interval (CI) for each cancer per log-odds increase in risk of VTE. Nominal P values (*P*), false discovery corrected P values (*FDR-P*) and heterogeneity P values for Cochrane’s Q statistic (*het-P*) are shown.

**Figure 2.**
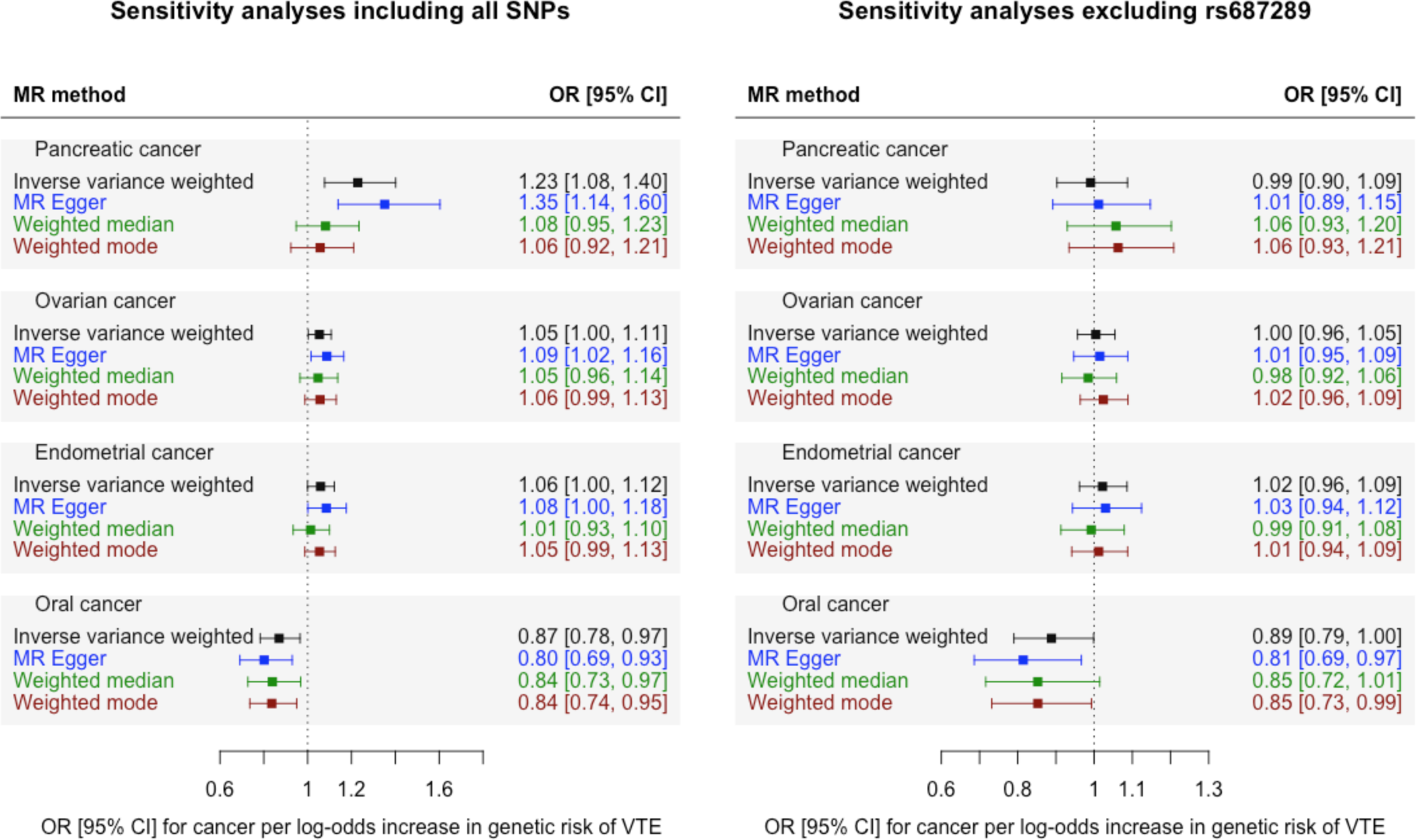
Mendelian randomisation sensitivity analyses of genetically-proxied risk of venous thromboembolism (VTE) as an exposure and four cancers (outcomes) which showed an association (*P*:<0.05) in the MR-IVW analysis (pancreatic, ovarian, endometrial and oral cancer). Left plot [A] shows sensitivity analyses including all SNPs. Right plot [B] shows sensitivity analyses with rs687289 removed. Results are represented as the odds ratio (OR) and 95% confidence interval (CI) for each cancer per log-odds increase in genetic risk of VTE.

There was weak evidence from the MR-IVW analysis for a small inverse association between genetic risk of VTE and risk of oral cancer (OR 0.87 [95% CI, 0.78-0.97], *P*=0.01, *FDR-P=*0.10). Sensitivity analyses showed consistent estimates of effect (**Figure 2, Supplementary Table S2**) with no indication of directional pleiotropy from the MR-Egger analysis (MR-Egger intercept 0.01, standard error 0.007). Heterogeneity tests indicated minimal heterogeneity (Cochrane’s Q statistic 28.7, *P*=1.00) and there were no obvious outliers on inspection of the funnel plots and leave-one-out plots (**Supplementary Figures S1-S2**).

We performed two additional sensitivity analyses: firstly using only 31 well replicated VTE-SNPs as instrumental variables (**Supplementary Tables S3 – S4, Supplementary Figure S4**), and secondly using all available VTE SNPs with no-Steiger filtering applied (**Supplementary Figure S5**). These results were similar to the primary analysis for all 18 cancers.

We examined the MR Wald ratios for the association between VTE risk proxied by either Factor V Leiden (rs6025) or Prothrombin G20210A (rs1799963), and cancer (**Figure 3, Supplementary Table S5**). Summary data for Factor V Leiden was available for all cancers. Summary data for Prothrombin G20210A was unavailable for 6 of 18 cancers (endometrial cancer, kidney cancer, lung cancer, marginal zone lymphoma, pancreatic cancer and prostate cancer). There was a very weak inverse association between VTE risk, proxied by Factor V Leiden, and colorectal cancer (OR 0.95 [95% CI, 0.90-1.00], *P=*0.04, *FDR-P* = 0.59). There were no associations between Factor V Leiden or Prothrombin G20210A and any other cancer.

**Figure 3.**
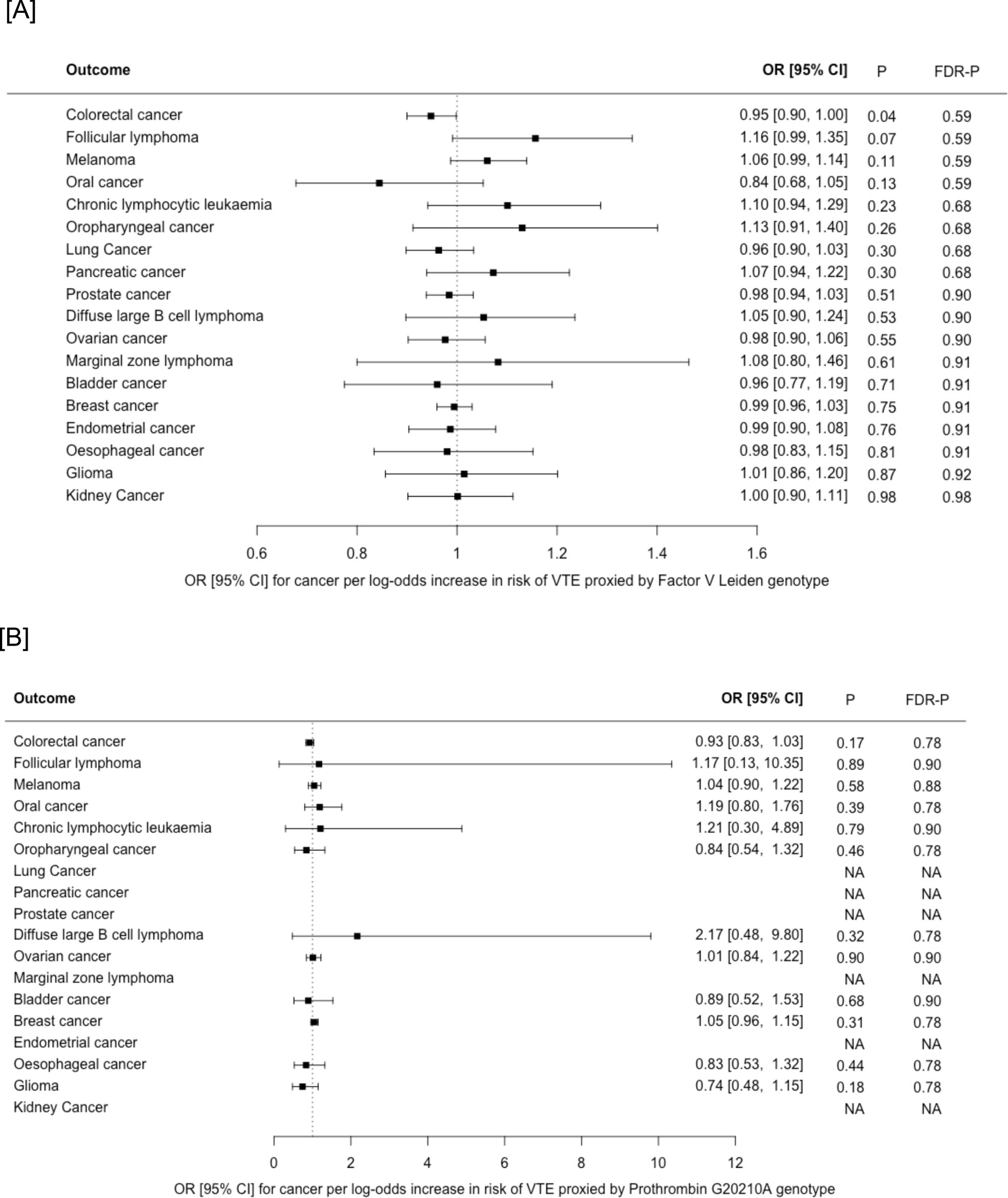
Mendelian randomisation Wald ratios for association between **[A]** venous thromboembolism (VTE), proxied by Factor V Leiden only), and risk of 18 cancers and **[B]** VTE, proxied by Prothrombin G20210A only, and risk of 12 cancers. Results are represented as the odds ratio (OR) and 95% confidence interval (CI) for each cancer per log-odds increase in genetic risk of VTE. Nominal P values (*P*) and false discovery corrected P values (*FDR-P*) are shown. NA indicates cancers for which the Prothrombin G20210A variant was not available in the GWAS summary data.

### ii) Mendelian randomisation analyses of the association between genetically-proxied risk of cancer and venous thromboembolism risk

We performed the MR analyses in the reverse direction, using genetically-proxied cancer-risk as an exposure and VTE as an outcome. The number of instrumental variables used for each cancer is shown in **Table 3**. Summary data for the SNPs used in each analysis is shown in **Supplementary Table S6**.

**Table 3.** Number of genetic instruments for each cancer used for Mendelian randomisation analysis, associated r^2^ (variance in cancer risk explained) and mean F statistic.

| Exposure GWAS | Cancer-risk SNPs<sup>a</sup> | SNPs unavailable<sup>a</sup> | SNPs excluded<sup>a</sup> | Cancer-SNPs used<sup>a</sup> | $r^2$ for cancer | Mean F statistic |
| --- | --- | --- | --- | --- | --- | --- |
| Breast cancer | 156 | 1 | 5 | 150 | 0.037 | 90 |
| Prostate cancer | 137 | 6 | 8 | 123 | 0.063 | 108 |
| Endometrial cancer | 16 | 0 | 0 | 16 | 0.013 | 44 |
| Colorectal cancer | 56 | 0 | 3 | 53 | 0.021 | 64 |
| Melanoma | 38 | 0 | 1 | 37 | 0.052 | 211 |
| Lung Cancer | 15 | 1 | 0 | 14 | 0.013 | 83 |
| Ovarian cancer | 12 | 0 | 1 | 11 | 0.010 | 71 |
| Kidney Cancer | 18 | 0 | 0 | 18 | 0.034 | 70 |
| Oesophageal cancer | 5 | 0 | 0 | 5 | 0.011 | 32 |
| Pancreatic cancer | 10 | 0 | 1 | 9 | 0.017 | 55 |
| Diffuse large B cell lymphoma | 4 | 0 | 0 | 4 | 0.012 | 27 |
| Chronic lymphocytic leukaemia | 8 | 0 | 0 | 8 | 0.048 | 70 |
| Follicular lymphoma | 2 | 0 | 0 | 2 | 0.032 | 48 |
| Oral cancer | 14 | 0 | 1 | 13 | 0.018 | 35 |
| Oropharyngeal cancer | 4 | 1 | 0 | 3 | 0.014 | 38 |
| Glioma | 5 | 0 | 0 | 5 | 0.028 | 80 |
| Marginal zone lymphoma | 1 | 0 | 0 | 1 | 0.009 | 10 |
| Bladder cancer | 2 | 0 | 0 | 2 | 0.006 | 31 |

<sup>a</sup> For each cancer exposure GWAS, 'Cancer-risk SNPs' refers to the total number of SNPs which could potentially be used as an IV for each cancer; 'SNPs unavailable' refers to the number of cancer-SNPs for which no direct correlate could be identified in the VTE GWAS study; 'SNPs excluded' refers to the number of SNPs which could not be harmonised due to coding-strand ambiguities or which were excluded after Steiger filtering. 'Cancer-SNPs used' refers to the final number of genetic instruments for cancer used in each analysis.

The MR-IVW analysis showed weak evidence for a small inverse association between genetically proxied risk of oropharyngeal cancer and risk of VTE (OR 0.93 [95% CI, 0.86-1.00], *P*=0.05, *FDR-P=*0.48, **Figure 4**). Only three SNPs were available as instrumental variables and these displayed significant heterogeneity in the MR-IVW analysis (Cochrane’s Q-statistic 11.0, *P=*0.004). This was reflected by the wide estimate of effect in the MR Egger analysis (OR 0.97 [95% CI, 0.61-1.52], *P=*0.90, MR Egger intercept: -0.02, intercept standard error: 0.08). Full results are shown in **Supplementary Table S7**. There were no other associations between genetically proxied risk of cancer and VTE in the primary analysis. However, a sensitivity analysis in which no Steiger-filtering was applied showed trends towards a possible association between genetically-proxied risk of pancreatic cancer and risk of VTE (OR 1.25, 95% CI 0.91 – 1.72, *P=*0.18, *FDR-P*=0.78) and also between genetically-proxied risk of ovarian cancer and risk of VTE (OR 1.18, 95% CI 0.78 – 1.79, *P*=0.42, *FDR-P*=0.87), **Supplementary Figure S6**. These associations (which were not seen in the primary analysis) resulted from the inclusion of an instrumental variable (rs687289 for pancreatic cancer, and rs115478735 for ovarian cancer) which had been excluded from the primary analysis by Steiger-filtering.

**Figure 4.**
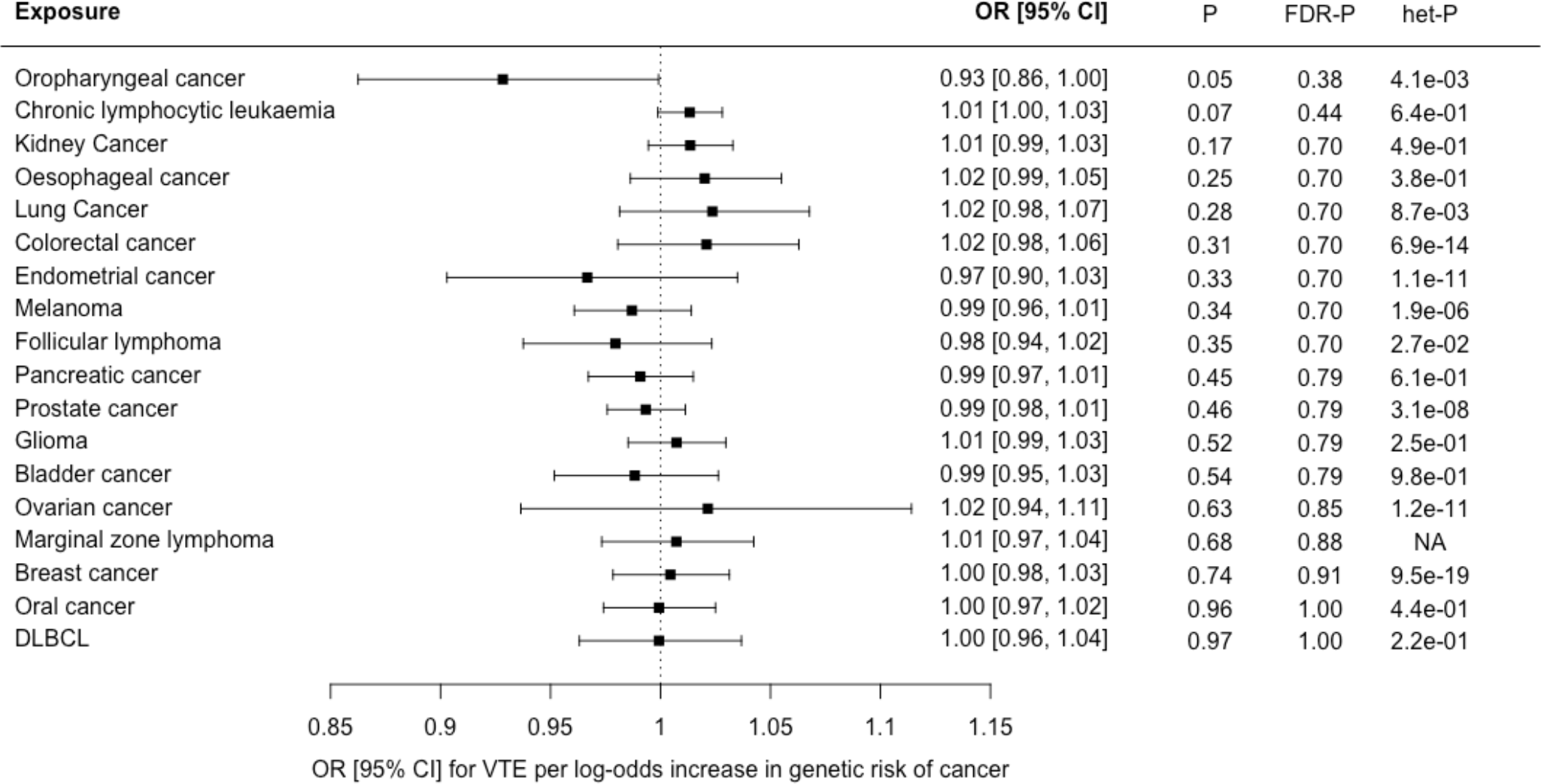
Forest plot showing estimates from Mendelian randomisation (MR) estimates of the effect of genetic risk of 18 cancers as exposures on venous thromboembolism (VTE), as an outcome. The MR inverse variance weighted estimate is shown for all cancers except marginal zone lymphoma, where the Wald ratio is shown as only a single instrumental variable was available. Results are represented as the odds ratio (OR) and 95% confidence interval (CI) for VTE per log-odds increase in genetic risk of each cancer. Nominal P values (*P*), false discovery corrected P values (*FDR-P*) and heterogeneity P values for Cochrane’s Q statistic (*het-P*) are shown.

### ii) Post-hoc power calculations

We calculated power for the MR analyses as described previously by Burgess *et al*.[45] Assuming a type 1 error rate < 0.05 and minimum effect estimate of OR≥1.5 for each outcome per log-odds increase in risk of exposure, there was >80% power to detect an association between genetic liability to VTE (as an exposure) and all cancers except marginal zone lymphoma and bladder cancer; there was >80% power to detect an association between genetic risk of all 18 cancers (as exposures) and VTE.

## Discussion

We observed an association between genetically-proxied risk of VTE and increased risk of pancreatic cancer. Pancreatic cancer has consistently been linked with very high rates of VTE in conventional epidemiological studies.[46–48] However, our MR sensitivity analyses indicated that the association between genetically-proxied risk of VTE and pancreatic cancer was largely driven by a single outlying SNP (rs687289). We also observed a weak association between VTE risk and risk of ovarian and endometrial cancer, which attenuated further towards the null in sensitivity analyses with rs687289 removed.

The variant rs687289 is found in intron 2 of the *ABO* blood group gene and the VTE-risk allele at this SNP is in high linkage disequilibrium with an allele which determines non-O blood group (rs8176719).[49] Non-O blood group is associated with increased risk of a range of phenotypes including cardiovascular disease and several cancers, including pancreatic and ovarian cancer.[50,51] One possibility is that the MR association between VTE and pancreatic cancer results from horizontal pleiotropy (i.e. rs687289 exerting an effect on both VTE and pancreatic cancer through independent biological pathways). People with non-O blood group have higher levels of von-Willebrand factor and LDL-cholesterol, both of which may potentially contribute to VTE.[49] The mechanism by which blood group affects cancer risk is unknown, although it is hypothesised that ABO antibodies interact with aberrant glycoproteins expressed on pancreatic tumour cells.[52] It is also plausible that the association between VTE and pancreatic cancer is mediated by ABO blood group. Further multivariable MR analyses were beyond the scope of this study but would be helpful in evaluating this hypothesis. Lastly, since the prevalence of different ABO blood groups varies geographically,[53] the associations driven by this SNP could indicate confounding by population stratification. Although all GWAS data were drawn from genetically-inferred European-ancestry participants, this encompasses a heterogenous group of people, defined by different GWAS using a variety of principal component clustering models. Therefore, there may be genetic drift between the cohorts included in the VTE and cancer studies.

There was weak evidence from both the MR-IVW and MR-sensitivity analyses that genetically increased risk of VTE was associated with a slightly reduced risk of oral cancer (MR-IVW OR 0.87 [95% CI, 0.78-0.97], *P*=0.01, *FDR*-*P=*0.10). Several previous studies have shown that for people presenting with VTE the risk of subsequent oral cancer diagnosis is relatively low compared to other cancers, an observation which adds credibility to our results.[54] A few small studies have previously described that SNPs in genes encoding two coagulation proteins, plasminogen activator inhibitor 1 (PAI-1) (rs1799889) and Factor XIII (rs5985) respectively, are associated with risk of oral squamous cell carcinoma.[55,56] Neither of these SNPs were identified as VTE-risk variants in the GWAS we extracted VTE risk SNPs from,[32] therefore these were not represented in our MR analysis. Their specific role in oral cancer carcinogenesis is unclear. Given the lack of a consistent biological mechanism to explain our MR finding, our result should be interpreted with caution.

We noted that Steiger filtering excluded more SNPs for cancers where the sample size of the outcome GWAS was much smaller than the VTE exposure GWAS. The Steiger test may be less reliable if there is a significant difference in measurement error between the exposure and outcome GWAS.[36] However, a sensitivity analysis in which all available VTE risk SNPs were used as IVs (no exclusions based on Steiger-filtering) found similar results for all cancers (**Supplementary Table S4**).

For the MR analysis in the cancer-VTE direction, which examined genetically-proxied risk of cancer as an exposure and VTE as an outcome, we found no clear evidence that genetic predisposition for any cancer was associated with an increased risk of VTE after correction for multiple testing. There are several caveats which should be considered in the interpretation of this result: This analysis estimates the impact of lifetime elevated genetic risk of cancer on risk of VTE and will not capture time-dependent causal associations which occur due to acute pathophysiological changes in the context of active or progressive malignancy. Secondly, if ‘cancer-associated’ VTE develops through biologically distinct mechanisms from VTE in the absence of cancer, a causal association may not be detected using this two sample MR approach in which the outcome VTE GWAS cohort was derived from a heterogenous case group with both provoked and unprovoked VTE (rather than a cancer-specific cohort).[32] Given these limitations, future MR studies using IVs which proxy time-dependent or intermediate exposure phenotypes may be helpful to explore the association between cancer and VTE.

### ii) Comparison with wider literature

Although associations between VTE and cancer have been rigorously examined by conventional epidemiological approaches,[2] to our knowledge, there are no published MR analyses examining the causal effect of genetic liability to VTE on cancer risk, and only one previous MR analysis examining the causal effect of genetic liability to cancer on VTE risk.[57] The authors of this study reported a trend towards reduced VTE risk in the context of genetic predisposition to melanoma (OR 0.89, 95% CI 0.82-0.97), and increased VTE risk in the context of genetic predisposition to non-hodgkin lymphoma (OR 1.20, 95% CI 1.00-1.44) and breast cancer (OR 1.09, 95% CI 1.00 – 1.20), although the evidence for these associations diminished after correction for multiple testing. In contrast, our study, which used data from GWAS with much larger case numbers for both VTE and each cancer, did not replicate these associations.

Several small case-control studies have applied regression analyses to examine whether carriers of single thrombophilia gene polymorphisms, including Factor V Leiden (rs6025) and prothrombin G20210A (rs1799963) are at increased risk of cancer.[58] Two groups previously reported that prothrombin G20210A was associated with an increased risk of gastrointestinal and colorectal cancer respectively.[39,41] In contrast, Vossen *et al*[40] found that heterozygous carriers of either prothrombin G20210A or Factor V Leiden had a reduced risk of colorectal cancer. Using an MR Wald ratio analysis, we also found weak evidence that the Factor V Leiden allele was associated with a slightly reduced risk of colorectal cancer (OR 0.94 [95% CI 0.89 - 1.00], *P*=0.04). Asymptomatic carriers of Factor V Leiden have been shown to have accelerated formation of activated protein C.[59] This enzyme has effects on endothelial barrier integrity and inflammation which appear to be independent of coagulation pathways.[60] Therefore, the inverse association between Factor V Leiden and colorectal cancer risk may result from a biological interaction which is independent of thrombosis. Alternatively, the result could reflect confounding by population stratification. Prothrombin G20210A genotype data was only available for 12 of the 18 cancers, however we did not find any associations between this variant and cancer as assessed by the MR Wald ratios.

## Conclusions

We present a bi-directional MR analysis examining the association between genetically-proxied risk of VTE and 18 different cancers, using summary data from large GWAS meta-analyses. Our findings do not support the hypothesis that VTE is a cause of cancer. Genetically-proxied risk of VTE was associated with an increased incidence of pancreatic cancer and slightly reduced incidence of oral cancer, but there was inadequate evidence to suggest a causal relationship. Further work is required to establish whether and how biological pathways involving ABO blood group contribute to epidemiological associations between VTE and pancreatic cancer. Additional mechanistic studies are required to elucidate causal relationships between active cancer and VTE, as well as the role of VTE in cancer progression.

## Ethics approval

This analysis used GWAS summary data only therefore no additional ethics approval was required. Approval was obtained by the original GWAS studies by their appropriate ethics review boards: see details in the source GWAS publications.

## Author contributions

NC, RL, AM and CT designed the study. HB, JF, TG, RG, MJ, DM, JM, RP, AP, NS, JS, FT, D-AT and CV contributed to the acquisition and analysis of primary data used for the analyses. NC performed the final analyses and drafted the manuscript with RL and PH. All authors critically revised and approved the final manuscript.

## Data availability statement

R scripts used for the analysis are available via GitHub [https://github.com/NaomiC-0/Mendelian-randomisation-analysis-of-VTE-and-Cancer]. Harmonised summary data for all SNPs included in this analysis are available in the Supplementary File. Full summary statistics are publicly available via: the Open GWAS database [https://gwas.mrcieu.ac.uk] for ovarian cancer (accession number: ieu-a-1120) and prostate cancer (accession number: ieu-b-85); the European Bioinformatics Institute GWAS Catalogue [https://www.ebi.ac.uk/gwas] for endometrial cancer (accession number: GCST006464), lung cancer (accession number: GCST004748) and oesophageal cancer (accession number: GCST003739); and the Breast Cancer Association Consortium [bcac.ccge.medschl.cam.ac.uk] for breast cancer. PanScan and PanC4 GWAS data are available through dbGAP (accession numbers phs000206.v5.p3 and phs000648.v1.p1, respectively). Application to the relevant GWAS consortium is required for full summary statistics for the remaining phenotypes.

## Supplementary data

See supplementary files

### Funding

This work was supported by the Wellcome Trust [grant number 225541/Z/22/Z (GW4 Clinical Academic Training Programme for Health Professionals) to NC] and Cancer Research UK [grant number C18281/A29019 to RL and PH]. JM was supported by the US National Cancer Institute [grant number 5U01CA257679-02]. D-A.T was supported by the EPIDEMIOM-VT Senior Chair from the University of Bordeaux initiative of excellence (IdEX). For the purpose of Open Access, the author has applied a CC BY public copyright licence to any Author Accepted Manuscript version arising from this submission.

Full funding acknowledgements for all contributing GWAS can be found in the source publications.

For the colorectal cancer GWAS: Genetics and Epidemiology of Colorectal Cancer Consortium (GECCO) was funded by: National Cancer Institute, National Institutes of Health, U.S. Department of Health and Human Services (R01 CA059045, U01 CA164930, R01 201407, R01 CA244588) and in part through the NIH/NCI Cancer Center Support Grant P30 CA015704. Scientific Computing Infrastructure at Fred Hutch was funded by ORIP grant S10OD028685. Colon CFR was funded by NCI/NIH award U01 CA167551. For acknowledgements and funding of specific contributing studies please see <u>full funding and acknowledgements</u>.

The breast cancer genome-wide association analyses for BCAC and CIMBA were supported by Cancer Research UK (PPRPGM-Nov20\100002, C1287/A10118, C1287/A16563, C1287/A10710, C12292/A20861, C12292/A11174, C1281/A12014, C5047/A8384, C5047/A15007, C5047/A10692, C8197/A16565) and the Gray Foundation, The National Institutes of Health (CA128978, X01HG007492-the DRIVE consortium), the PERSPECTIVE project supported by the Government of Canada through Genome Canada and the Canadian Institutes of Health Research (grant GPH-129344) and the Ministère de l’Économie, Science et Innovation du Québec through Genome Québec and the PSRSIIRI-701 grant, the Quebec Breast Cancer Foundation, the European Community’s Seventh Framework Programme under grant agreement n° 223175 (HEALTH-F2-2009-223175) (COGS), the European Union’s Horizon 2020 Research and Innovation Programme (634935 and 633784), the Post-Cancer GWAS initiative (U19 CA148537, CA148065 and CA148112 - the GAME-ON initiative), the Department of Defence (W81XWH-10-1-0341), the Canadian Institutes of Health Research (CIHR) for the CIHR Team in Familial Risks of Breast Cancer (CRN-87521), the Komen Foundation for the Cure, the Breast Cancer Research Foundation and the Ovarian Cancer Research Fund. All studies and funders are listed in Zhang H et al (Nat Genet, 2020).

The International Lung Cancer Consortium (ILCCO) was supported by grant U19 CA203654

Funding for the kidney cancer GWAS was provided by the US National Institutes of Health (NIH), National Cancer Institute (U01CA155309) for those studies coordinated by IARC and by the intramural research program of the National Cancer Institute, US NIH, for those studies coordinated by the NCI.

For the melanoma GWAS: The MelaNostrum Consortium was funded by the intramural Research Program of the National Institutes of Health (NIH), National Cancer Institute (NCI), Division of Cancer Epidemiology and Genetics and by the participating institutes in Italy, Spain, and Greece. The GenoMEL study was funded by the European Commission under the 6th Framework Programme (contract no. LSHC-CT-2006-018702), by Cancer Research UK Programme Awards (C588/A4994 and C588/A10589), by a Cancer Research UK Project Grant (C8216/A6129) and by a grant from the US National Institutes of Health (NIH; CA83115). This research was also supported by the intramural Research Program of the NIH, National Cancer Institute (NCI), Division of Cancer Epidemiology and Genetics, and by grants from the National Health and Medical Research Council (NHMRC) of Australia. For acknowledgements and funding of specific contributing studies please see the source publication <LANDI 2020>

## Disclaimer

Where authors are identified as personnel of the International Agency for Research on Cancer/World Health Organization, the authors alone are responsible for the views expressed in this article, and they do not necessarily represent the decisions, policy, or views of the International Agency for Research on Cancer/World Health Organization. Where authors are identified as affiliated with National Heart, Lung and Blood Institute or the National Institute of Health, the views expressed in this manuscript are those of the authors and do not necessarily represent the views of the National Heart, Lung and Blood Institute or the National Institute of Health

## Acknowledgements

This work has been conducted using GWAS summary data (including publicly available data) from separate GWAS consortia. We are grateful to all the consortia and their participants. These include the: Breast Cancer Association Consortium (BCAC), Barrett’s and Esophageal Adenocarcinoma Consortium (BEACON), Esophageal Adenocarcinoma Genetics Consortium (EAGLE), Endometrial Cancer Association Consortium (ECAC), Genetics and Epidemiology of Colorectal Cancer Consortium (GECCO), Genetic Epidemiology of Glioma International Consortium, International Lung Cancer Consortium (ILCCO), International Lymphoma Epidemiology Consortium (InterLymph), INVENT-VTE and MVP consortia, MelaNosrtum Consortium, GenoMEL Consortium, Oncoarray Consortium, Ovarian Cancer Association Consortium (OCAC), Pancreatic Cancer Cohort Consortium (PanScan) and Pancreatic Cancer Case-Control Consortium (PanC4), Prostate Cancer Association Group to Investigate Cancer Associated Alterations in the Genome Consortium (PRACTICAL), the Nijmegen Bladder Cancer Study (NBCS) and Nijmegen Biomedical Study. This work used the computational facilities of the Advanced Computing Research Centre, University of Bristol – http://www.bristol.ac.uk/acrc

