## Supplementary Figures for "Causal relationships between risk of venous thromboembolism and 18 cancers: a bidirectional Mendelian randomisation analysis"

Supplementary Figures: Contents

Supplementary Figure S1:
Leave one out plots for selected Mendelian randomisation analyses of venous thromboembolism (exposure) and pancreatic, ovarian, endometrial and oral cancer.

Supplementary Figure S2: Funnel plots for selected Mendelian randomisation analyses of venous thromboembolism (exposure) and pancreatic, ovarian, endometrial and oral cancer

Supplementary Figure S3: Single SNP plots for selected Mendelian randomisation analyses of venous thromboembolism (exposure) and pancreatic, ovarian, endometrial and oral cancer

Supplementary Figure S4: Forest plot for Mendelian randomisation inverse variance weighted analyses of venous thromboembolism (exposure, with instrumental variables restricted to replicated SNPs only) and 18 cancers

Supplementary Figure S5: Forest plot for Mendelian randomisation inverse variance weighted analyses of venous thromboembolism (exposure, with instrumental variables including all available SNPs – no Steiger-filtering applied) and 18 cancers

Supplementary Figure S6: Forest plot for Mendelian randomisation analyses of 18 cancers (exposures, with instrumental variables including all available SNPs – no Steiger-filtering applied) and venous thromboembolism.

Figure S1A

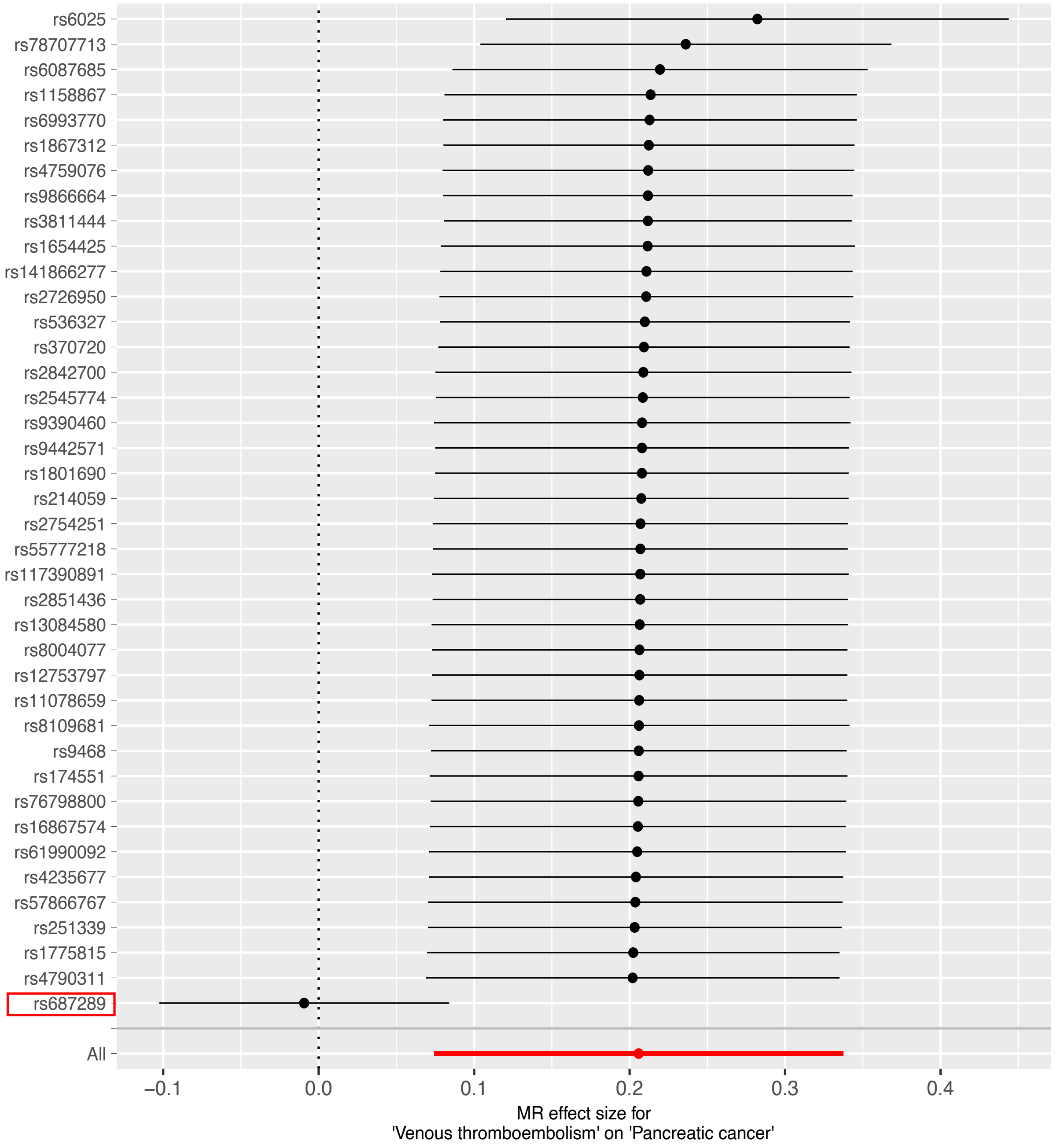


Supplementary Figure S1 [A – D]:
Leave one out plots for selected MR analyses of VTE (exposure) and [A] pancreatic, [B] ovarian, [C] endometrial and [D] oral cancer. The x-axis shows the MR-IVW effect estimates (log-OR) after sequential removal of each SNP shown on the y-axis. The variant rs687289 which proxies non-O blood group is highlighted.

Figure S1B

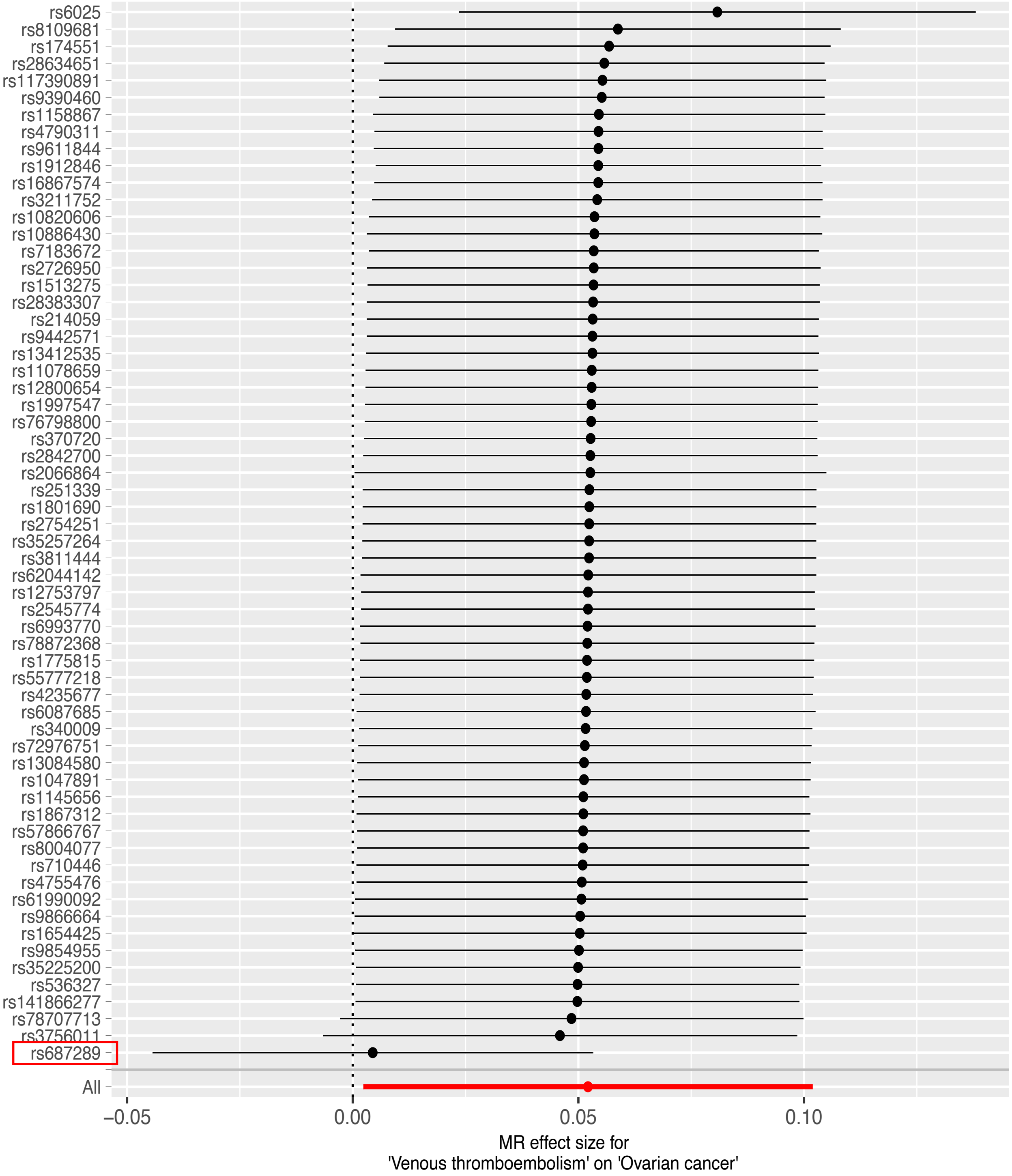
rs6025
rs8109681
rs174551
rs28634651
rs117390891
rs9390460
rs1158867
rs4790311
rs9611844
rs1912846
rs16867574
rs3211752
rs10820606
rs10886430
rs7183672
rs2726950
rs1513275
rs28383307
rs214059
rs9442571
rs13412535
rs11078659
rs12800654
rs1997547
rs76798800
rs370720
rs2842700
rs2066864
rs251339
rs1801690
rs2754251
rs35257264
rs3811444
rs62044142
rs12753797
rs2545774
rs6993770
rs78872368
rs1775815
rs55777218
rs4235677
rs6087685
rs340009
rs72976751
rs13084580
rs1047891
rs1145656
rs1867312
rs57866767
rs8004077
rs710446
rs4755476
rs61990092
rs9866664
rs1654425
rs9854955
rs35225200
rs536327
rs141866277
rs78707713
rs3756011
rs687289
All
−0.05
0.00
0.05
0.10
MR effect size for
'Venous thromboembolism' on 'Ovarian cancer'

Figure S1C

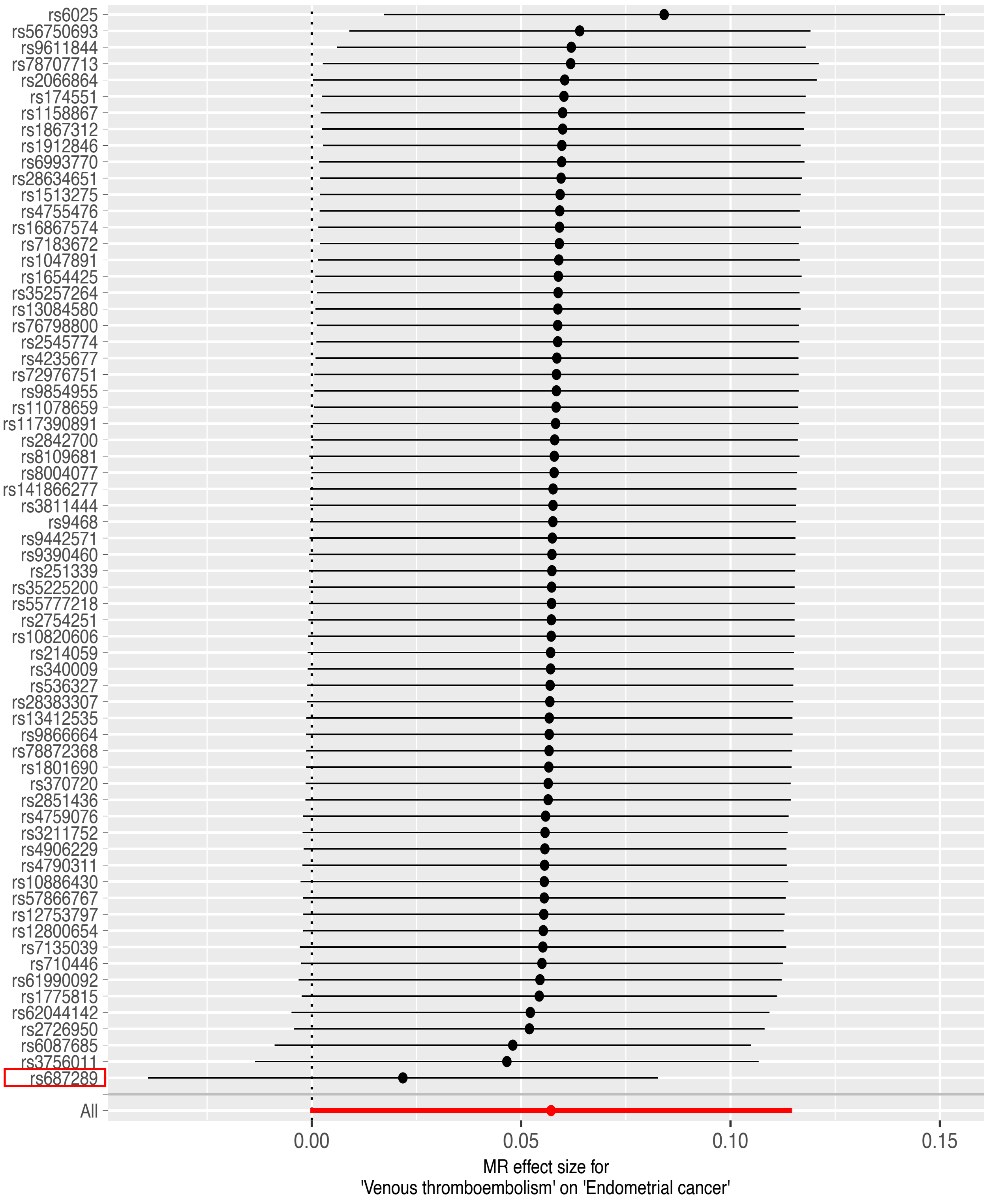
rs6025
rs56750693
rs9611844
rs78707713
rs2066864
rs174551
rs1158867
rs1867312
rs1912846
rs6993770
rs28634651
rs1513275
rs4755476
rs16867574
rs7183672
rs1047891
rs1654425
rs35257264
rs13084580
rs76798800
rs2545774
rs4235677
rs72976751
rs9854955
rs11078659
rs117390891
rs2842700
rs8109681
rs8004077
rs141866277
rs3811444
rs9468
rs9442571
rs9390460
rs251339
rs35225200
rs55777218
rs2754251
rs10820606
rs214059
rs340009
rs536327
rs28383307
rs13412535
rs9866664
rs78872368
rs1801690
rs370720
rs2851436
rs4759076
rs3211752
rs4906229
rs4790311
rs10886430
rs57866767
rs12753797
rs12800654
rs7135039
rs710446
rs61990092
rs1775815
rs62044142
rs2726950
rs6087685
rs3756011
rs687289
All
0.00
0.05
0.10
0.15
MR effect size for
'Venous thromboembolism' on 'Endometrial cancer'

Figure S1D

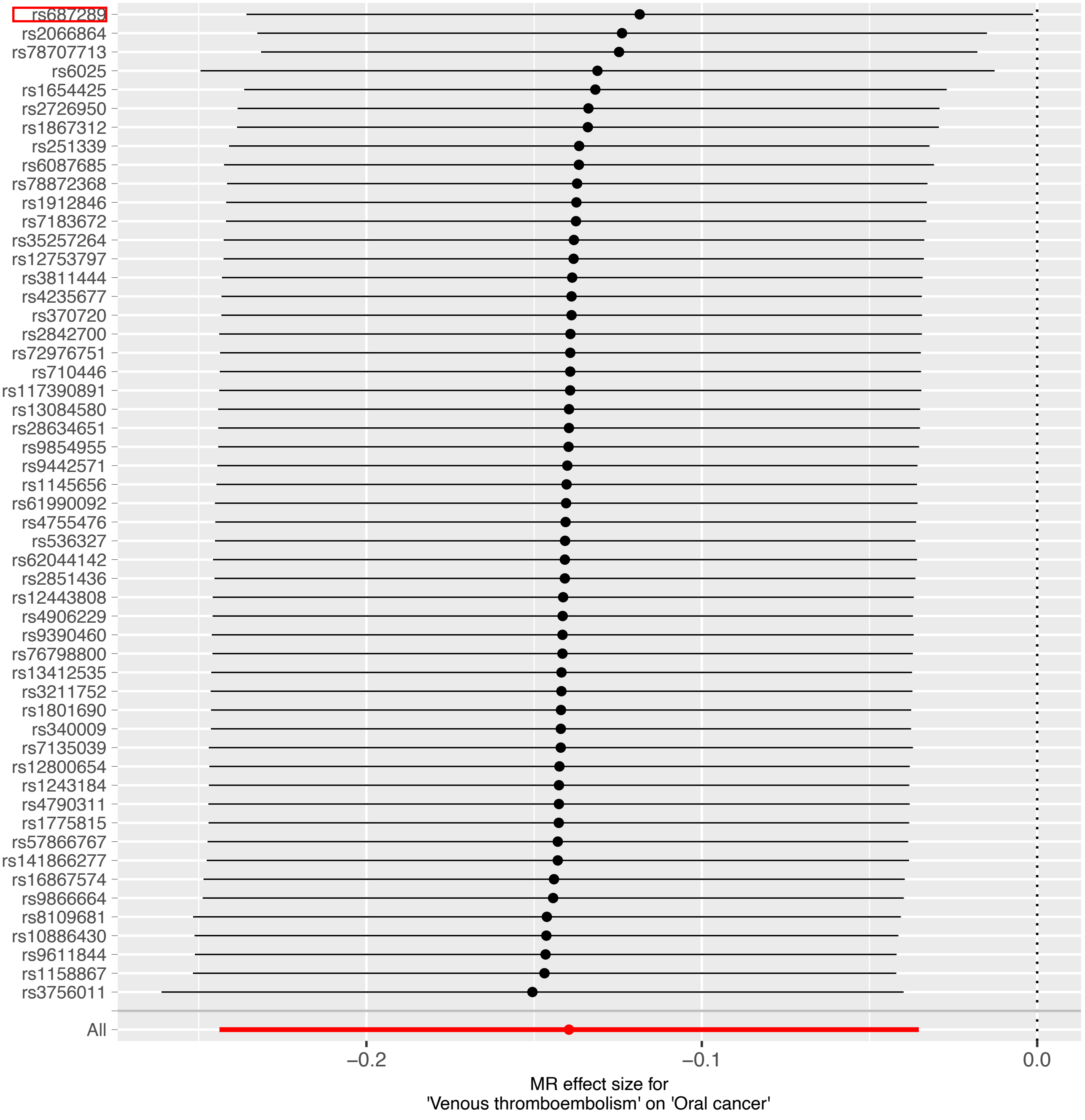
rs687289
rs2066864
rs78707713
rs6025
rs1654425
rs2726950
rs1867312
rs251339
rs6087685
rs78872368
rs1912846
rs7183672
rs35257264
rs12753797
rs3811444
rs4235677
rs370720
rs2842700
rs72976751
rs710446
rs117390891
rs13084580
rs28634651
rs9854955
rs9442571
rs1145656
rs61990092
rs4755476
rs536327
rs62044142
rs2851436
rs12443808
rs4906229
rs9390460
rs76798800
rs13412535
rs3211752
rs1801690
rs340009
rs7135039
rs12800654
rs1243184
rs4790311
rs1775815
rs57866767
rs141866277
rs16867574
rs9866664
rs8109681
rs10886430
rs9611844
rs1158867
rs3756011
All
−0.2
−0.1
0.0
MR effect size for
'Venous thromboembolism' on 'Oral cancer'

Figure S2A

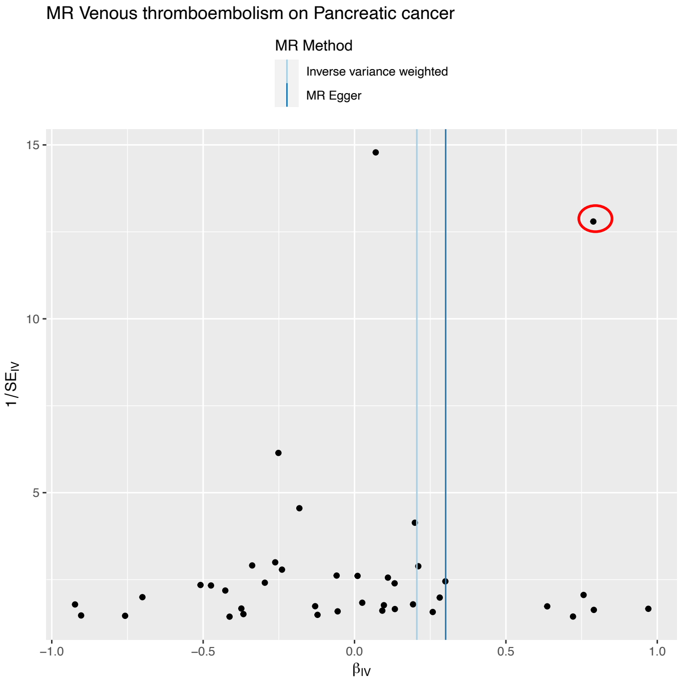


Figure S2B

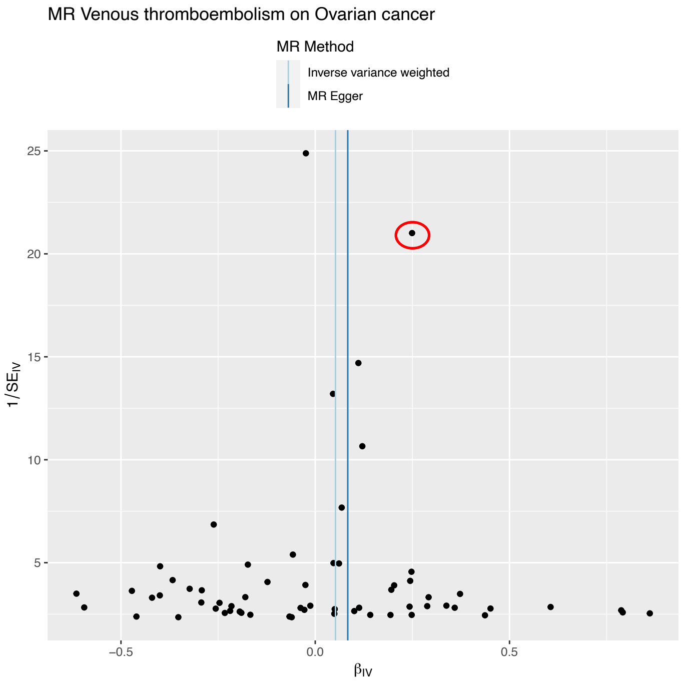


Figure S2C

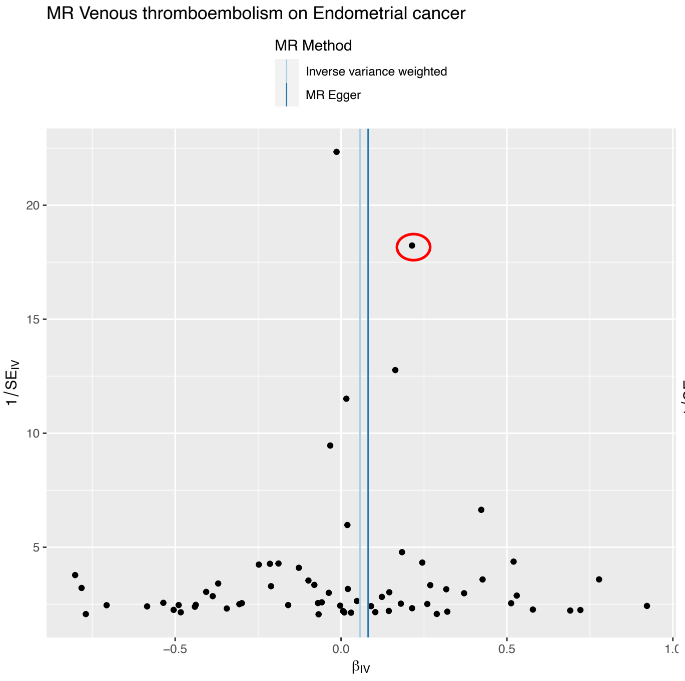


Figure S2D

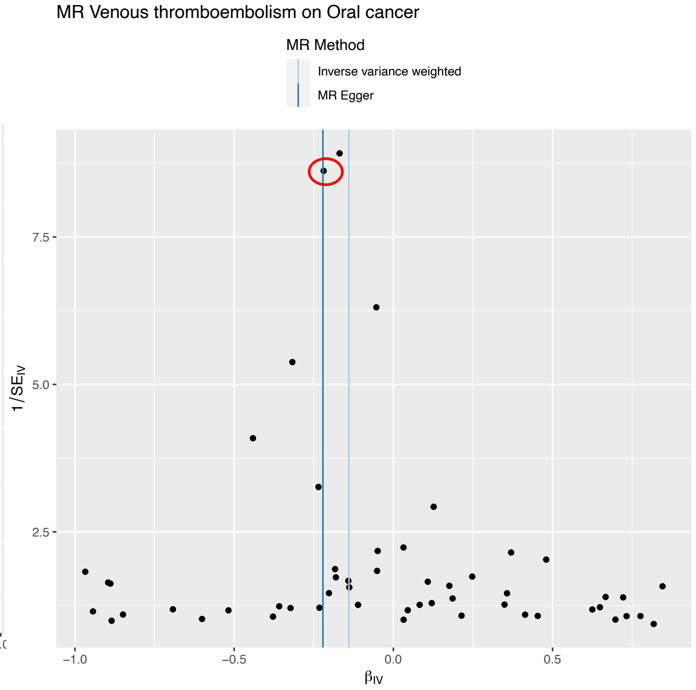


Supplementary Figure S2:
Funnel plots for selected MR analyses of VTE (exposure) and [A] pancreatic, [B] ovarian, [C] endometrial and [D] oral cancer. The x-axis shows the MR Wald ratio effect estimate (log-OR) for each SNP (IV); the y-axis shows the inverse standard error of the effect estimate. The variant rs687289 which proxies non-O blood group is circled.

Figure S3A

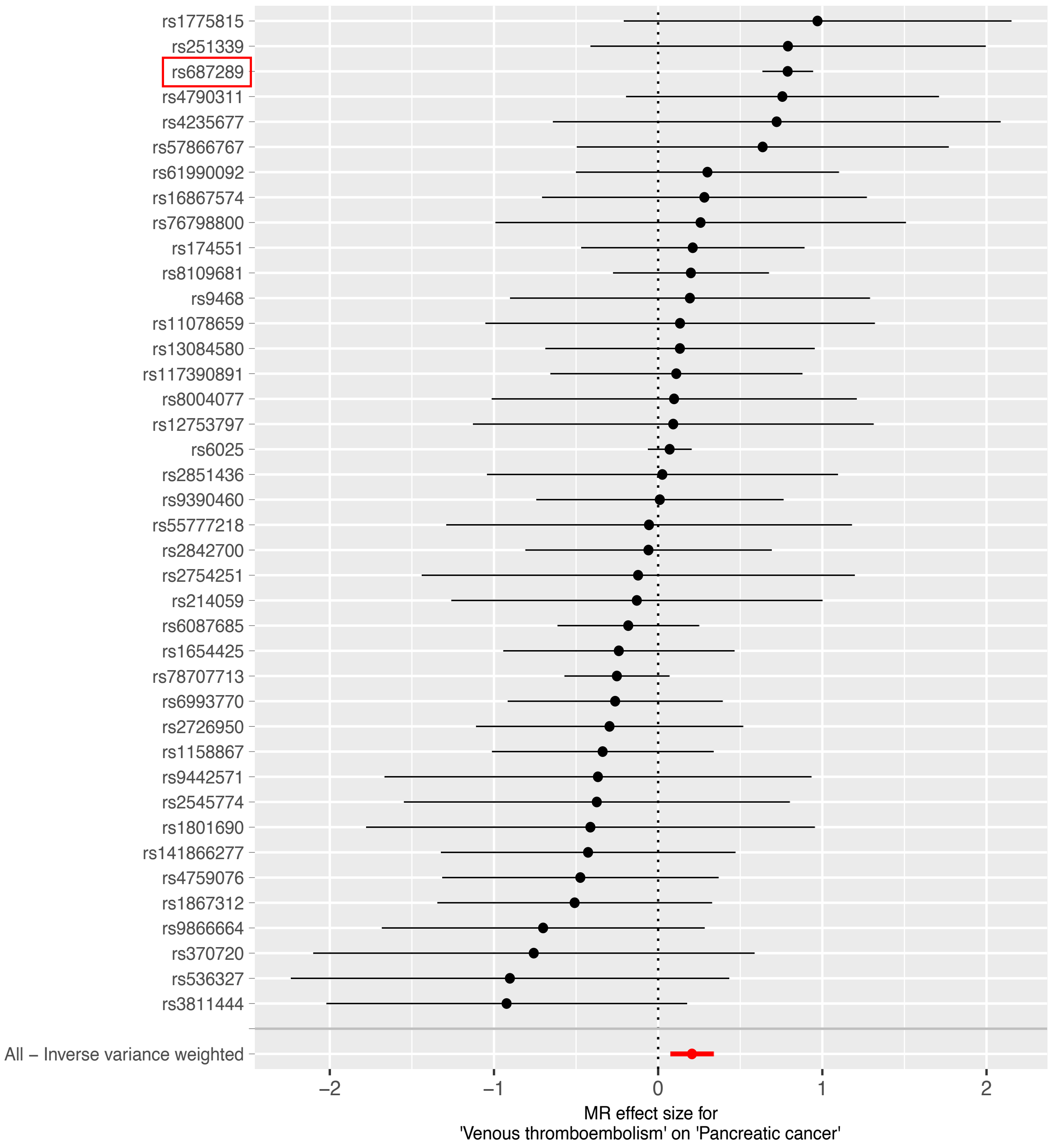


Supplementary Figure S3 [A-D]:
Single SNP plots for selected MR analyses of VTE (exposure) and [A] pancreatic, [B] ovarian, [C] endometrial and [D] oral cancer. The x-axis shows the MR Wald ratio effect estimate (expressed as log-OR) for each SNP shown on the y-axis. The variant rs687289 which proxies non-O blood group is highlighted in red.

Figure S3B

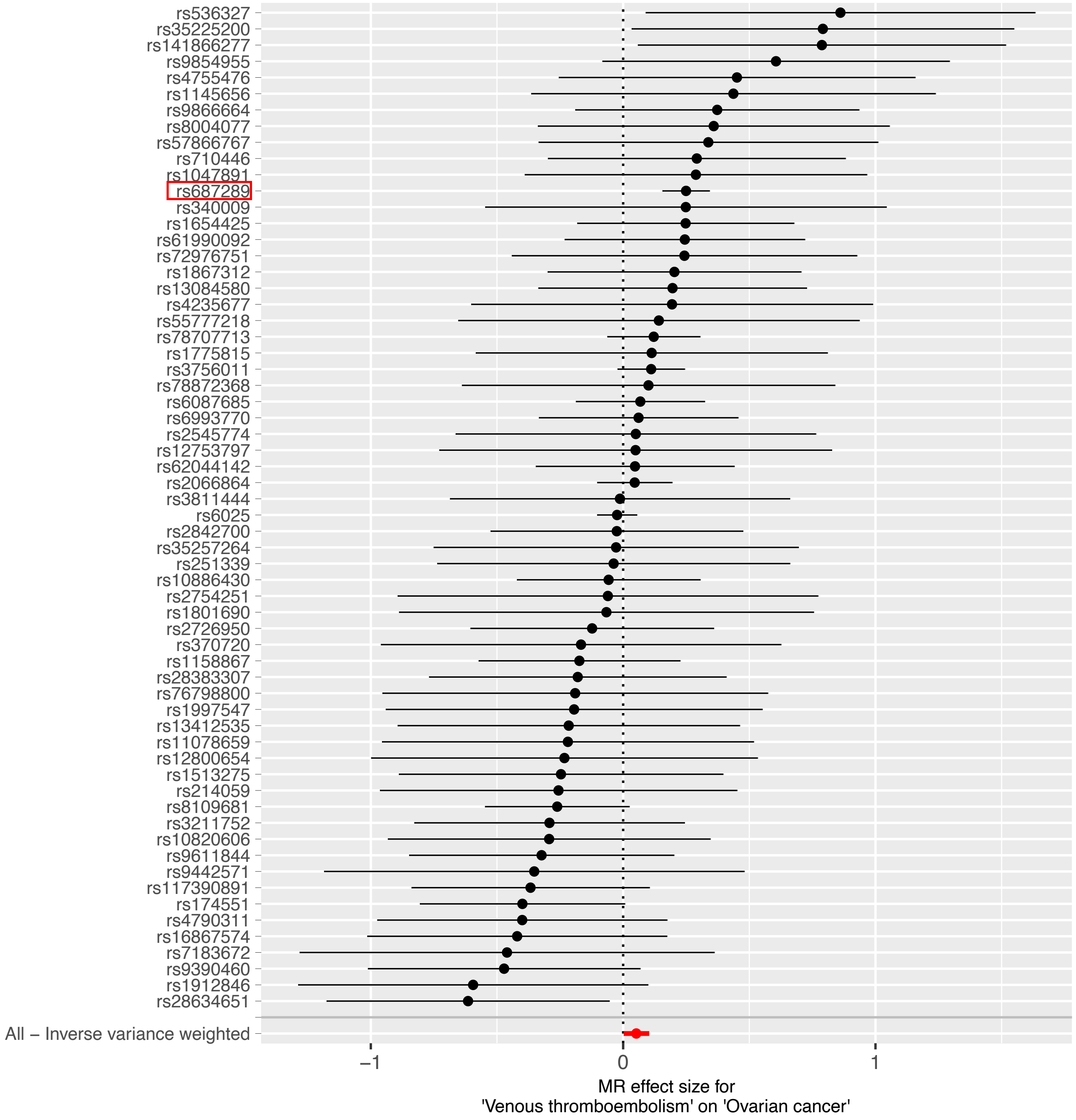
rs536327
rs35225200
rs141866277
rs9854955
rs4755476
rs1145656
rs9866664
rs8004077
rs57866767
rs710446
rs1047891
rs687289
rs340009
rs1654425
rs61990092
rs72976751
rs1867312
rs13084580
rs4235677
rs55777218
rs78707713
rs1775815
rs3756011
rs78872368
rs6087685
rs6993770
rs2545774
rs12753797
rs62044142
rs2066864
rs3811444
rs6025
rs2842700
rs35257264
rs251339
rs10886430
rs2754251
rs1801690
rs2726950
rs370720
rs1158867
rs28383307
rs76798800
rs1997547
rs13412535
rs11078659
rs12800654
rs1513275
rs214059
rs8109681
rs3211752
rs10820606
rs9611844
rs9442571
rs117390891
rs174551
rs4790311
rs16867574
rs7183672
rs9390460
rs1912846
rs28634651
All – Inverse variance weighted
–1
0
1
MR effect size for
'Venous thromboembolism' on 'Ovarian cancer'

Figure S3C

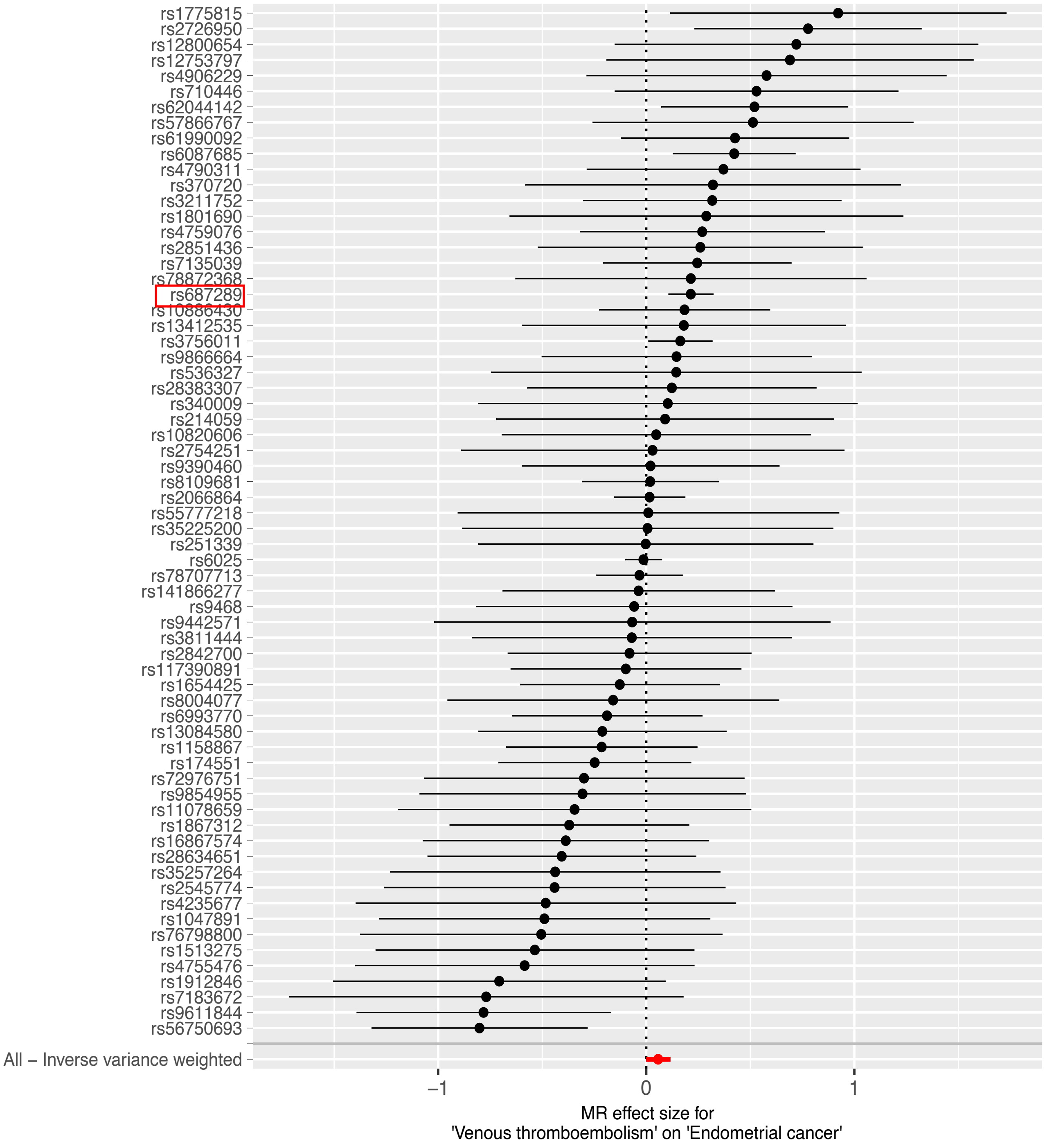

rs1775815
rs2726950
rs12800654
rs12753797
rs4906229
rs710446
rs62044142
rs57866767
rs61990092
rs6087685
rs4790311
rs370720
rs3211752
rs1801690
rs4759076
rs2851436
rs7135039
rs78872368
rs687289
rs10886430
rs13412535
rs3756011
rs9866664
rs536327
rs28383307
rs340009
rs214059
rs10820606
rs2754251
rs9390460
rs8109681
rs2066864
rs55777218
rs35225200
rs251339
rs6025
rs78707713
rs141866277
rs9468
rs9442571
rs3811444
rs2842700
rs117390891
rs1654425
rs8004077
rs6993770
rs13084580
rs1158867
rs174551
rs72976751
rs9854955
rs11078659
rs1867312
rs16867574
rs28634651
rs35257264
rs2545774
rs4235677
rs1047891
rs76798800
rs1513275
rs4755476
rs1912846
rs7183672
rs9611844
rs56750693
All – Inverse variance weighted
−1
0
1
MR effect size for
'Venous thromboembolism' on 'Endometrial cancer'

Figure S3D

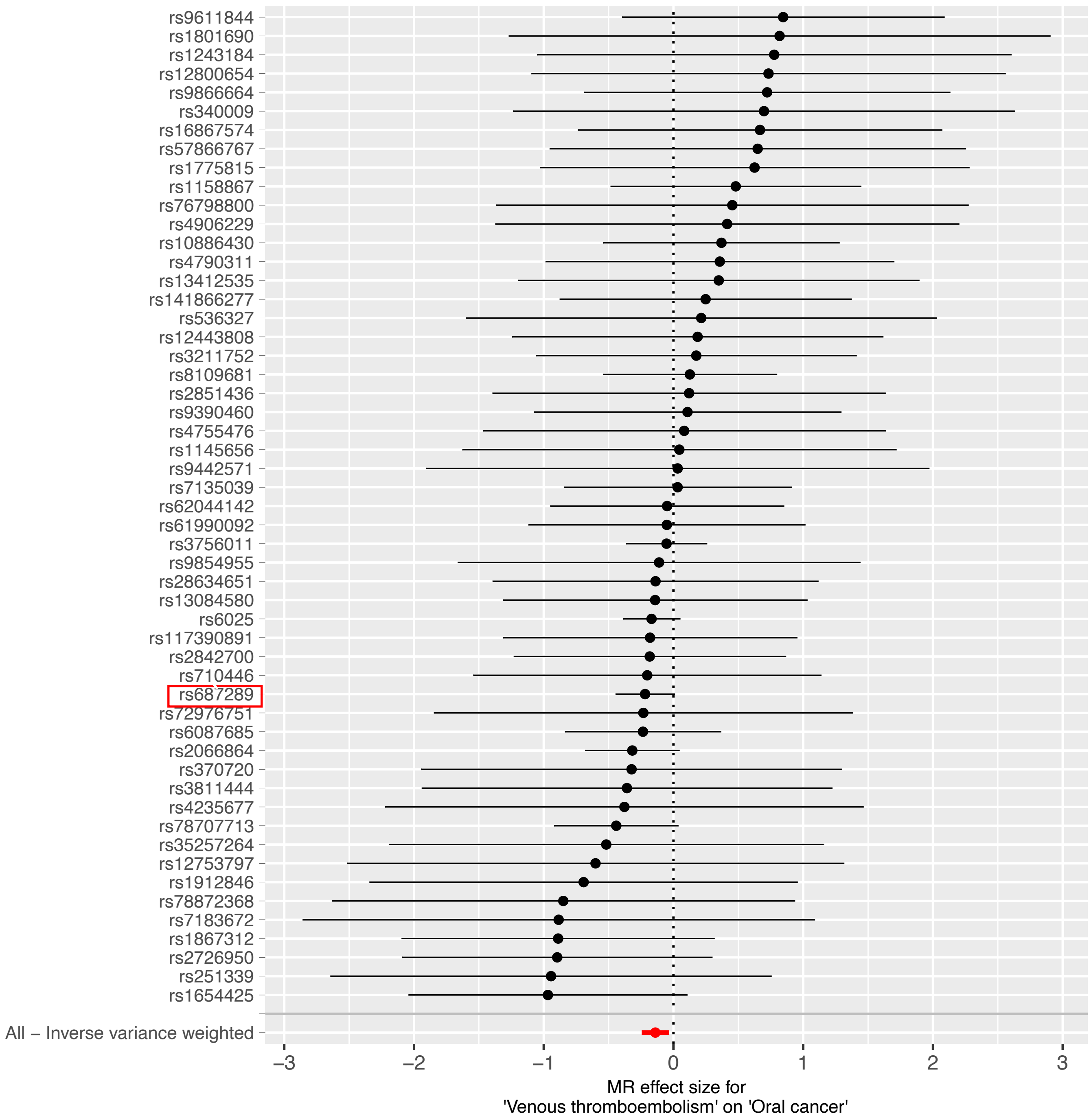
rs9611844
rs1801690
rs1243184
rs12800654
rs9866664
rs340009
rs16867574
rs57866767
rs1775815
rs1158867
rs76798800
rs4906229
rs10886430
rs4790311
rs13412535
rs141866277
rs536327
rs12443808
rs3211752
rs8109681
rs2851436
rs9390460
rs4755476
rs1145656
rs9442571
rs7135039
rs62044142
rs61990092
rs3756011
rs9854955
rs28634651
rs13084580
rs6025
rs117390891
rs2842700
rs710446
rs687289
rs72976751
rs6087685
rs2066864
rs370720
rs3811444
rs4235677
rs78707713
rs35257264
rs12753797
rs1912846
rs78872368
rs7183672
rs1867312
rs2726950
rs251339
rs1654425
All – Inverse variance weighted
−3
−2
−1
0
1
2
3
MR effect size for
'Venous thromboembolism' on 'Oral cancer'

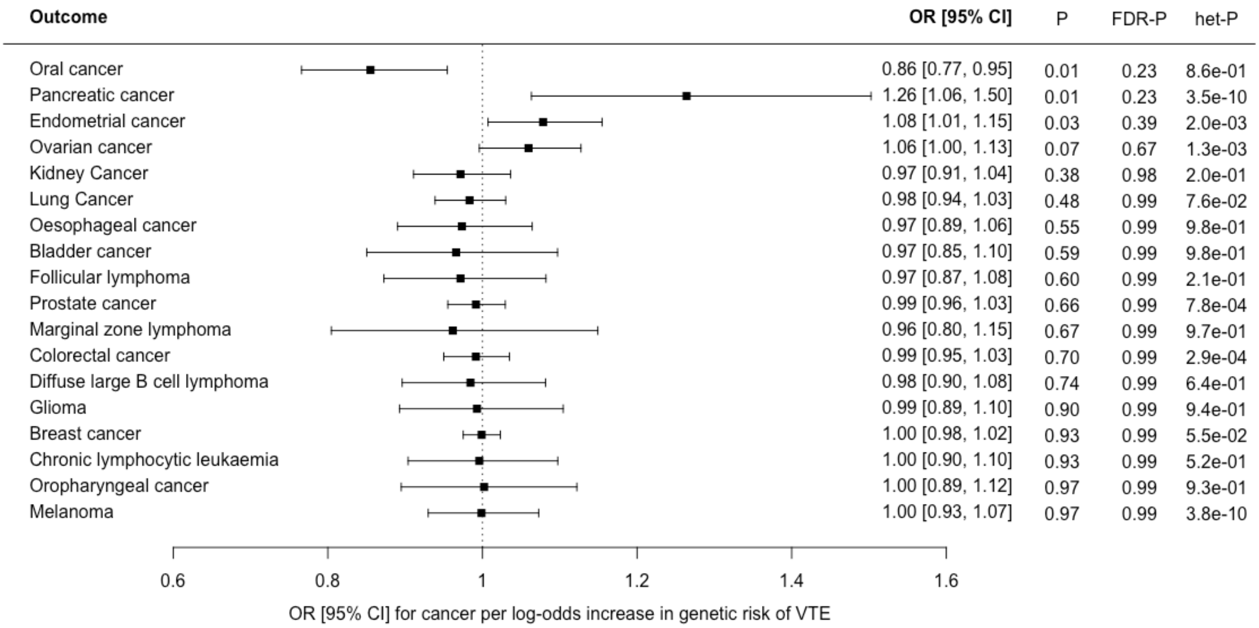


Supplementary Figure S4: Forest plot showing estimates from Mendelian randomisation inverse variance weighted estimates for VTE as an exposure, with instrumental variables restricted to replicated SNPs only, and 18 cancers as outcomes. Results are represented as the odds ratio (OR) and 95% confidence interval (CI) for each cancer per log-odds increase in genetic risk of each VTE. Nominal P values (*P*), false discovery corrected P values (*FDR-P*) and heterogeneity P values for Cochrane's Q statistic (*het-P*) are shown.

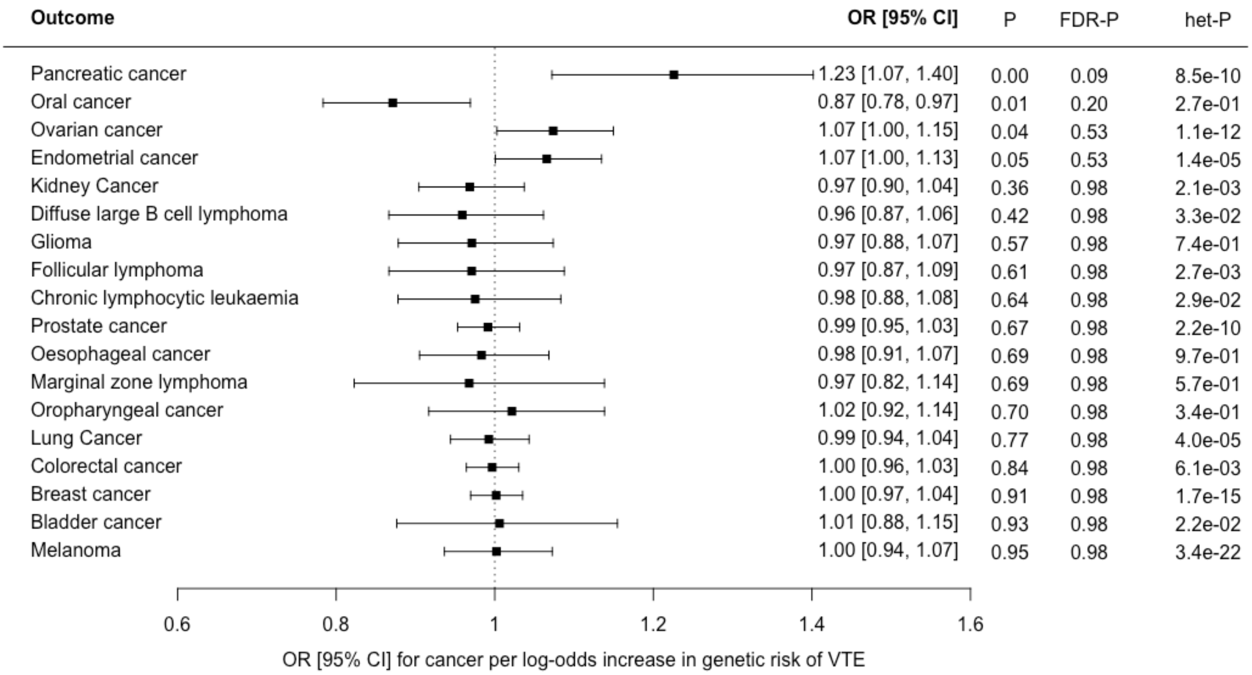


Supplementary Figure S5: Forest plot showing estimates from Mendelian randomisation inverse variance weighted estimates for VTE as an exposure, with instrumental variables including all available VTE SNPs (no Steiger-filtering applied), and 18 cancers as outcomes. Results are represented as the odds ratio (OR) and 95% confidence interval (CI) for each cancer per log-odds increase in genetic risk of each VTE. Nominal P values (*P*), false discovery corrected P values (*FDR-P*) and heterogeneity P values for Cochrane's Q statistic (*het-P*) are shown

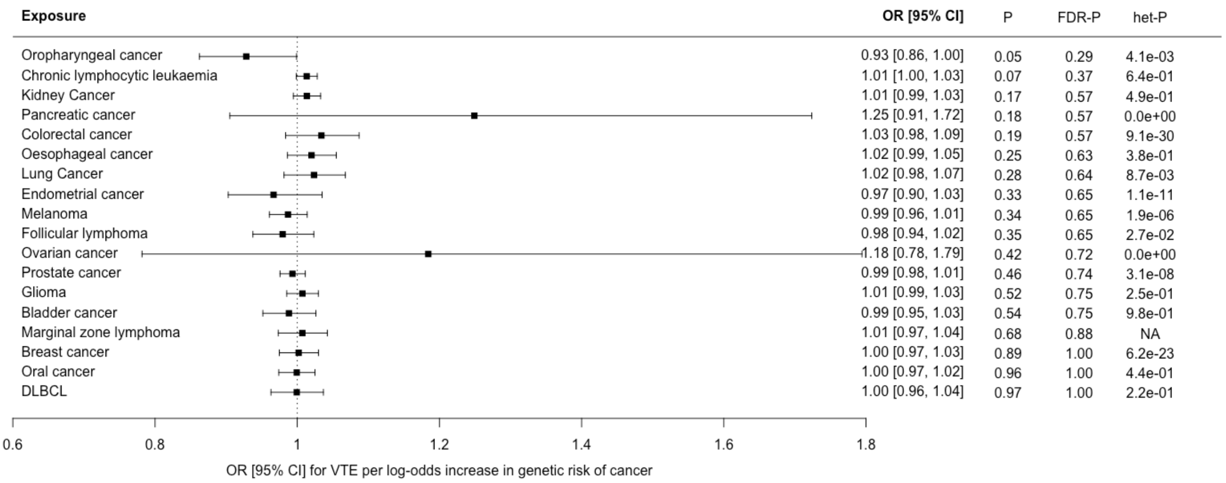


Supplementary Figure S6: Forest plot showing estimates from Mendelian randomisation (MR) estimates for 18 cancers as exposures, with instrumental variables including all available cancer-risk SNPs (no Steiger-filtering applied), and VTE as an outcome. The MR inverse variance weighted estimate is shown for all cancers except Marginal zone lymphoma, where the Wald ratio is shown as only a single instrumental variable was available. Results are represented as the odds ratio (OR) and 95% confidence interval (CI) for VTE per log-odds increase in genetic risk of each cancer. Nominal P values (*P*), false discovery corrected P values (*FDR-P*) and heterogeneity P values for Cochrane's Q statistic (*het-P*) are shown.
